# Double-Bowtie Filter Design for Pediatric Spectral CT Imaging

**DOI:** 10.64898/2026.01.15.26344121

**Authors:** Yinglin Ge, Olivia F. Sandvold, Roland Proksa, Amy E. Perkins, Thomas Koehler, Kevin M. Brown, Yannan Jin, Heiner Daerr, Ravindra M. Manjeshwar, Peter B. Noël

**Affiliations:** Department of Radiology, Perelman School of Medicine, University of Pennsylvania, Philadelphia, USA; Department of Bioengineering, University of Pennsylvania, Philadelphia, USA; Philips Healthcare, Cleveland, USA; Philips Innovative Technologies, Hamburg, Germany

**Keywords:** computed tomography (CT), quantitative imaging, spectral CT, DECT, kVp-switching, pediatric imaging, bowtie filter

## Abstract

**Purpose:** To develop and evaluate a novel double bowtie filter integrating a K-edge material layer with a conventional Teflon filter for pediatric spectral computed tomography (CT). The proposed design aims to enhance spectral signal-to-noise ratio (SNR) and spectral separation while maintaining radiation dose levels suitable for pediatric imaging.

**Methods:** A simulation framework was set up and used to model a rapid kVp-switching CT system operating at 70/110 kVp with realistic tube power and geometry constraints. Pediatric phantoms of three sizes (100– 200 mm anterior–posterior width) were used to evaluate performance. Five accessible and safe filter materials-gadolinium (Gd), holmium (Ho), erbium (Er), silver (Ag), and tin (Sn)-were tested in combination with a Teflon bowtie. System performance was quantified using virtual monoenergetic image (VMI) SNR at 40 keV and 70 keV, and the area under the monoenergetic SNR curve (AUMC) as a comprehensive spectral image quality metric. Dose consistency with a traditional Teflon bowtie reference was enforced.

**Results:** The Teflon + Gd configuration achieved the highest performance, improving AUMC by 47.5 % on average and up to 56 % for the largest phantom. VMI SNR increased by approximately 49 % at 40 keV and 42 % at 70 keV.

**Conclusions:** The double-bowtie concept substantially enhances spectral performance. The Teflon + Gd design provides a manufacturable, pediatric-optimized solution adaptable to kVp-switching and other spectral CT architectures, offering improved diagnostic quality at low dose levels.

## 1. INTRODUCTION

Unlike conventional CT, which merges all X-ray energies into a single signal, spectral CT distinguishes between different energy levels, allowing it to differentiate materials that appear similar on conventional scans^1,2^. Spectral data enables the creation of material maps and provides access to quantitative biomarkers^3^, supporting more accurate diagnostic screening, disease staging, and treatment monitoring. Spectral CT also enables the reconstruction of virtual monoenergetic images (VMI), which can enhance iodine contrast visibility, improve attenuation consistency, and reduce artifacts^2,4–6^. These capabilities offer significant clinical advantages across a wide range of applications. While minimizing radiation exposure remains paramount in pediatric imaging, CT continues to be the gold standard for children because of its high speed and superior spatial resolution. Spectral CT has demonstrated clinical advantages in pediatric diagnostics compared to conventional CT, including improved lesion characterization and artifact reduction^7^.

There are several spectral CT instrumentations, including source-based and detector-based techniques. Source-based systems contain dual-source dual-energy, single-source kilovoltage switching (kVp-switching)^8^, and split-beam systems such as twin-beam dual-energy^9^, which use additional filtration^10,11^. Detector-based spectral CT includes dual-layer and photon-counting^12–14^, each offering distinct performance in energy resolution and spectral separation ^15^. In spectral CT, spectral separation refers to the system’s ability to distinguish between low-and high-energy X-ray photons. This separation is essential for accurate and stable material decomposition and contrast agent quantification. By increasing spectral separation, the reliability and precision of measurements are improved, resulting in better image quality for all results and the possibility of a lower radiation dose^16^. Low tube voltages are commonly used in pediatric CT protocols to reduce radiation dose^17^. However, this results in reduced spectral separation in spectral CT^18,19^.

K-edge materials, which exhibit sharp increases in X-ray absorption at specific photon energies, offer a potential solution. By selectively attenuating parts of the spectrum, they can enhance spectral separation (**Figure 1**). Incorporating a K-edge filter between the X-ray source and the patient generates a spectrally shaped beam, improving energy discrimination and potentially restoring or improving image quality even at lower tube voltages ^20,21^. This concept is in use in commercial sources-based spectral CT system when applying a stronger filtration for the high tube voltage spectrum compared to the low tube voltage spectrum to reduce the spectral overlap.

**Figure 1.**
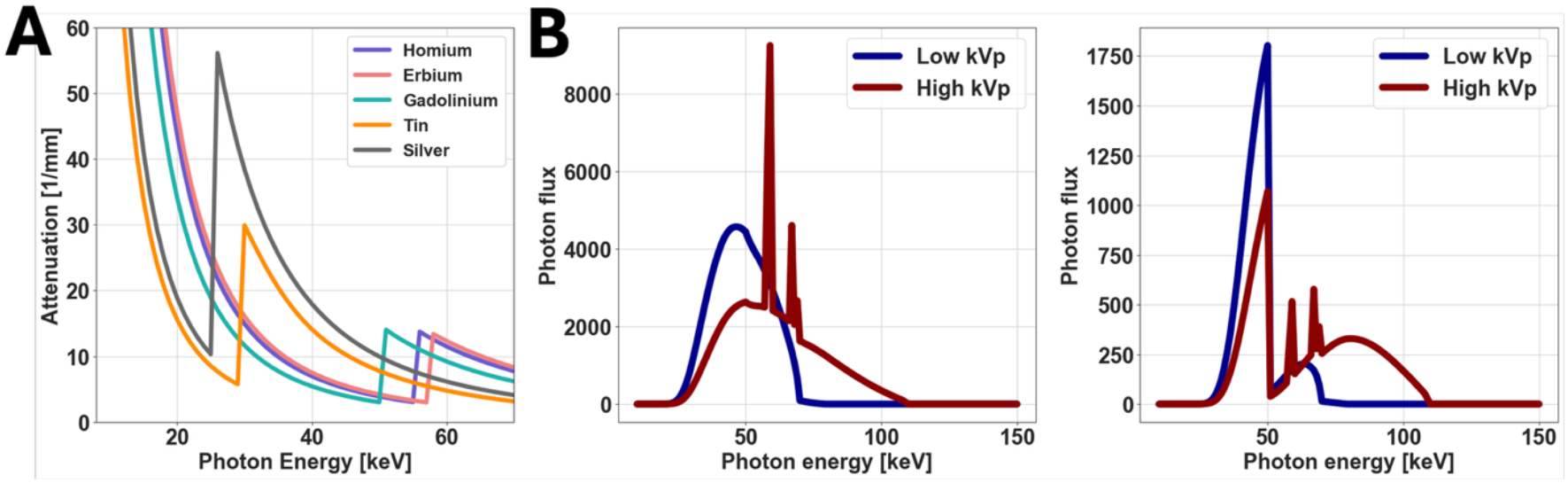
Attenuation of K-edge materials and spectra before and after K-edge filtration. (**A**) Attenuation curve of selected K-edge materials. (B) *Left:* Low kVp (70 kVp) and high kVp (110 kVp) spectra of simulated kVp-switching systems. *Right:* Spectra attenuated by 0.3 mm gadolinium.

In CT technology, the bowtie filter remains a critical hardware component. Its fan-angle-dependent shape is designed to reduce unnecessary radiation to the patient’s periphery, helping to optimize dose distribution. Additionally, bowtie filters help equalize the X-ray signal across the detector array, reducing the detector’s dynamic range requirements^22^ and reducing pulse pile-up effects for photon-counting detectors (PCDs)^23^. Teflon (CF_2_) is widely used as a bowtie filter material due to its attenuation properties, which closely resemble those of water, making it approximately tissue-equivalent^22^. However, thicker Teflon filters can contribute to increased scatter, and geometrical constraints of the CT gantry may limit the feasible thickness. To address these concerns, some systems use aluminum (Al) as an alternative material^24^. While aluminum allows for a more compact design, its higher atomic number (Z) makes it less tissue-equivalent. Besides different materials, bowtie filters can be tailored for specific anatomical regions to compensate for variations in tissue composition and shape^25,26^. Non-traditional designs, such as dynamic bowtie filters^27–30^, further aim to adapt the filter shape across different projection angles for improved patient matching. However, these approaches focus largely on optimizing material choice and geometry, with little integration of spectral CT requirements^31^. Moreover, there has been limited effort dedicated to optimizing these designs specifically for pediatric patients, the population most vulnerable to radiation exposure.

Although CT is widely used in pediatric imaging, most commercial CT systems are optimized for adults^32^. To address this challenge, we propose a novel double bowtie prefiltration design that pairs a conventional Teflon bowtie filter with a K-edge filter, delivering enhanced image quality for pediatric spectral CT imaging. The K-edge material selectively shapes the X-ray spectrum, enhancing spectral separation, while its strong attenuation properties allow for a thinner Teflon bowtie, thereby reducing scatter. Meanwhile, the bowtie filter continues to fulfill its essential roles in normalizing detector signals and decreasing unnecessary patient dose. In our previous work, this combined design demonstrated improved spectral performance in simulations using a dual-layer CT system under parallel beam geometry^33^ for a limited number of K-edge materials^34^. In this study, we expand upon that foundation by exploring additional K-edge materials and optimizing the double bowtie design in fan-beam geometry for a kVp-switching CT system. We hypothesize that integrating a K-edge layer within a conventional bowtie filter can enhance spectral separation and SNR without increasing radiation dose.

## 2. METHODS

### 2.1 System simulation

A Python simulation was designed to model a CT system comprised of clinical-grade components. X-ray beamlets were modeled from a prototype rapid kVp-switching tube (vMRC 800, Philips Healthcare) with a maximum tube power limit of 80 kW. For kVp-switching configurations, the voltage rise time was 44 µs and the fall time was 34 µs. The X-ray spectra emitted from the anode was generated using spekCalc software^35^ with photon energy from 10 keV to 150 keV. The spectra after filtration, and the realistic detector responses were then simulated (**Figure 2**). The material and thickness of the inherit filtration of the X-ray tube were incorporated into the model. Photon scattering and absorption data of materials were sourced from the National Institute of Standards and Technology (NIST) XCOM dataset^36^. The CT gantry geometry was also modeled based on the clinically feasible CT system.

**Figure 2.**
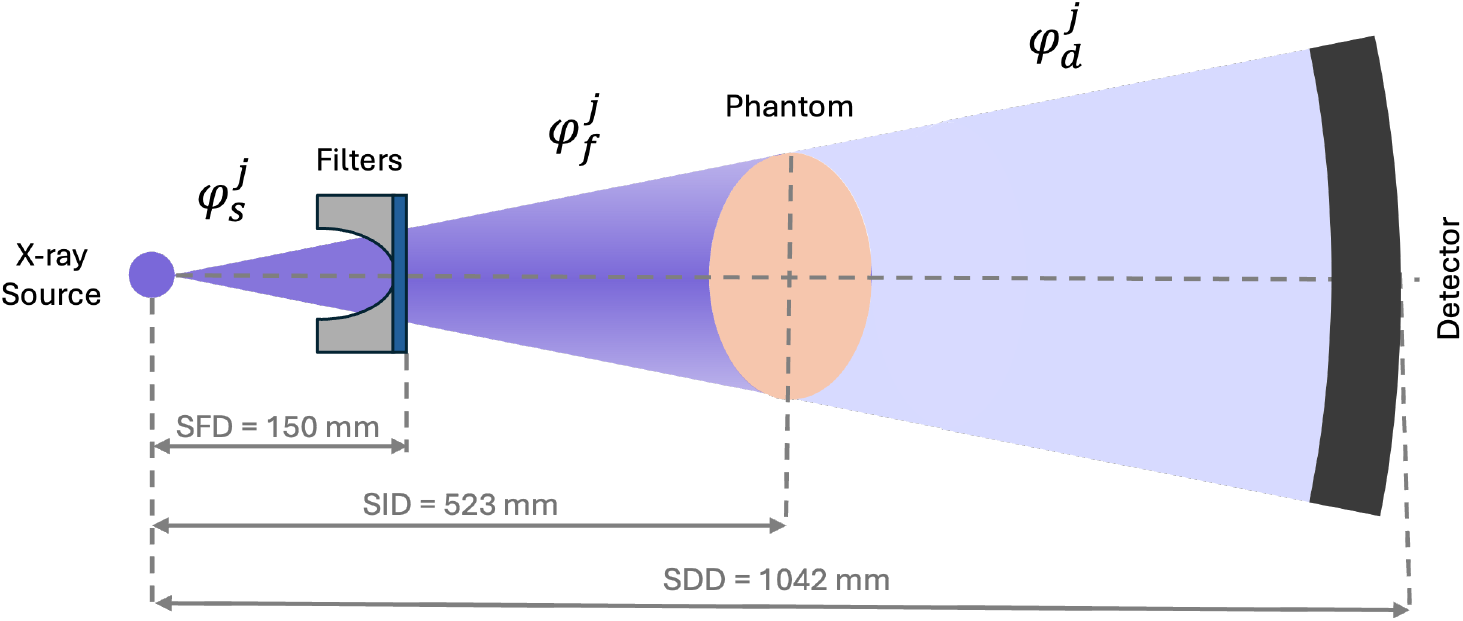
System sketch. For each ray *j*, the initial x-ray spectrum was denoted as 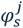, the spectrum after passing the bowtie filter was denoted as 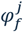, and the spectrum after passing the phantom and would be collected by the detector was denoted as 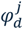, respective. The cosine effect (right) was applied for all *j*^th^ x-rays outside of 0 degrees in the simulated fan-beam. The source-to-filter distance (SFD) was 150 mm, the source-to-isocenter distance (SID) was 523 mm, and the source-to-detector distance (SDD) was 1042 mm.

The tube voltage waveform of the kVp-switching system is shown in Figure 3. The tube current changed with the changing tube voltage, higher tube current shortens the voltage transition duration during ramp-down. In this study, X-ray exposure was simulated at tube voltage pairs of 110 kVp (high-kVp, *φ*_110_)and 70 kVp (low-kVp, *φ*_70_) with an average tube current of 100 mA. The total rotation time was 1000 ms, with a duty cycle of 10/90, indicating the percentage of time allocated to high- and low-kVp spectra, respectively. A total of 2400 readings (1200 for each voltage) were acquired per rotation. These settings defined the reference system, mimicking a dose level comparable to standard pediatric abdominal or pediatric chest imaging protocols.

**Figure 3.**
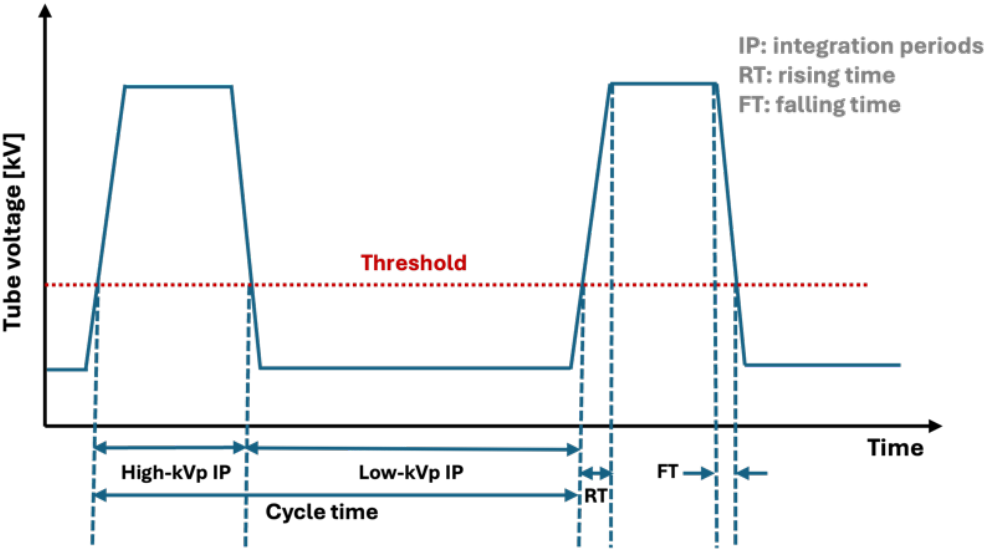
Simplified waveform of tube voltage in the kVp-switching system. One cycle time consists of a high-kVp integration period (IP) and a low-kVp IP, which are separated by a threshold voltage. For illustrative purposes, the duty cycle is set to 33/67.

### 2.2 Phantom design

To model pediatric patients of varying sizes, three elliptical water phantoms were simulated. The selected phantom sizes represented a broad range of pediatric age groups, with anteroposterior widths of 100 mm, 150 mm, and 200 mm, and transverse widths of 150 mm, 232 mm, and 313 mm. The size corresponded to the average abdominal dimensions of newborns, 8.7-year-olds, and 17.5-year-olds, respectively ^37^.

### 2.3 System evaluation

#### 2.3.1 Detector SNR

The detector signal signal-to-noise ratio (SNR) was calculated from the spectra *φ*_d_(*V, E*) received at the detector, where we assumed the photon shot noise followed the Poisson distribution. In an energy*-* integrating detector, each detected photon contributes an amount of signal response proportional to its energy. The signal from the impinging spectrum *φ*_110,70_ can be represented by total photon energy. The noise is the standard deviation in the distribution, so the detector SNR can be written as

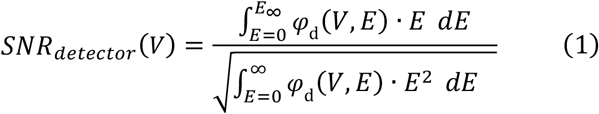

where *V* is the tube voltage. In our simulation, energies from 20 to 150 keV are calculated.

#### 2.3.2 VMI SNR

In this simulation, the Cramér-Rao lower bound of variance (CRLB) was used to estimate the noise on the photoelectric effect and Compton scatter two-basis decomposition^38^. The attenuation coefficient of a material was assumed to be the linear combination of these two bases functions. Spectral CT reconstruction was based on the estimation of line integrals *A*_*k*_ along a ray *C* from two spectra measurements^39^, where *k* represents photoelectric effect (pe) or Compton scatter (cs). *a*_*k*_ is spatial density map, often referred to as the material-specific coefficient.

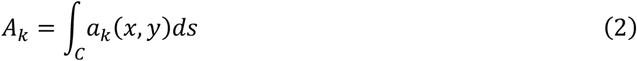

The energy dependent attenuation caused by this material can be written as

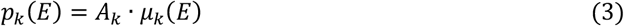

where *μ*_*k*_ is the energy-dependent attenuation coefficient of the material *k*. The two spectra received, *φ*_d_(110 *kV, E*) and *φ*_d_(70 *kV, E*), are assumed to be independent signals. Each measurement *M*_*j*_ was modeled as a Gaussian random variable representing the total energy deposited in the *j*^th^ acquisition, with *m*_*j*_ denoting its observed value, for *j* = 1, …, *N*. Let *θ*_*j*_ and 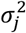 be the corresponding mean and variance of the Gaussian distribution measured at tube voltage *V*_*j*_ (Equations 4-5, where the material line integrals got another index *j*).

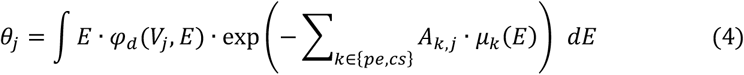

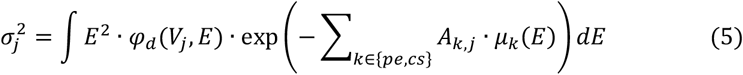

Then the probability density *ρ*(*m*_1_, …, *m*_*N*_) of *M*_*j*_ can be calculated as follows:

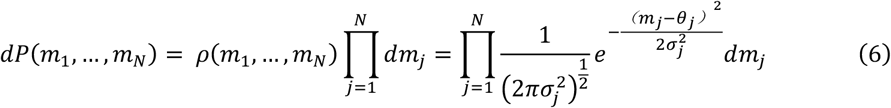

The negative log-likelihood can therefore be written as:

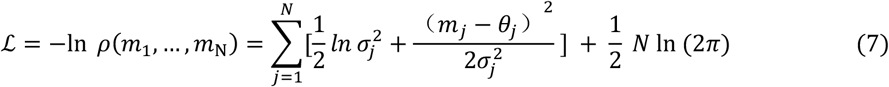

As mentioned before, ℒ is dependent on the attenuation of photoelectric effect and Compton scatter, thus also dependent on *A*_*pe*_ and *A*_*cs*_. The Fisher information matrix, ℱ(*θ*), can be derived by taking the expectation of the partial derivation of likelihood function with respect to *A*_*pe*_ and *A*_*cs*_,

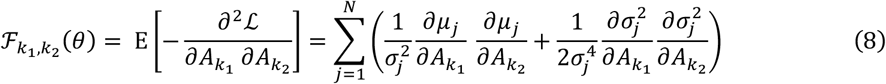

We can now obtain the Cramér–Rao inequality^40^ for the variance of any unbiased estimator of the parameters *A*_*pe*_ and *A*_*cs*_:

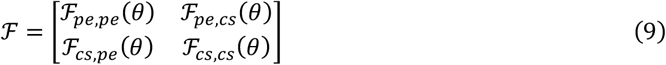

Where the determinant of the **ℱ is:

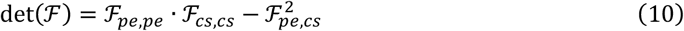

Then the separate CRLB bound for each basis can be written as:

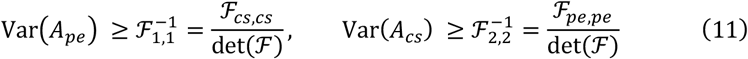

To assess the performance of material decomposition at specific energies, we compute the local SNR at a monoenergetic energy *E* based on the propagated uncertainty from the CRLB. The variance of each energy level, denoted as 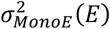, can be calculated from the CRLB covariance matrix cov [ℱ]^-1^:

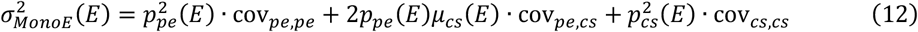

The approximate noise at each VMI level could then be defined as the square root of the variance, as 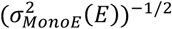.

In spectral CT, 40 keV VMI is commonly used for enhancing the contrast of iodine contrast agents and soft tissue^41,42^. Additionally, 70 keV VMI is better representative of conventional image^43^ and yields relative lower image noise with a higher iodine contrast-to-noise ratio (CNR)^42,44^. Given the clinical utility and importance, SNRs of VMI 40 keV and VMI 70 keV are calculated.

#### 2.3.3 AUMC calculation

To assess overall spectral image quality, a noise metric termed the area under the mono-energy curve (AUMC) is introduced. AUMC is calculated as the integral of the inverse of the mono *E* CRLB standard deviation estimates from 40 to 140 keV (**Figure 4**).

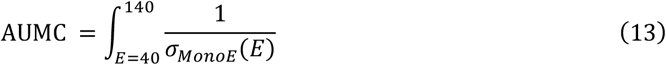

**Figure 4.**
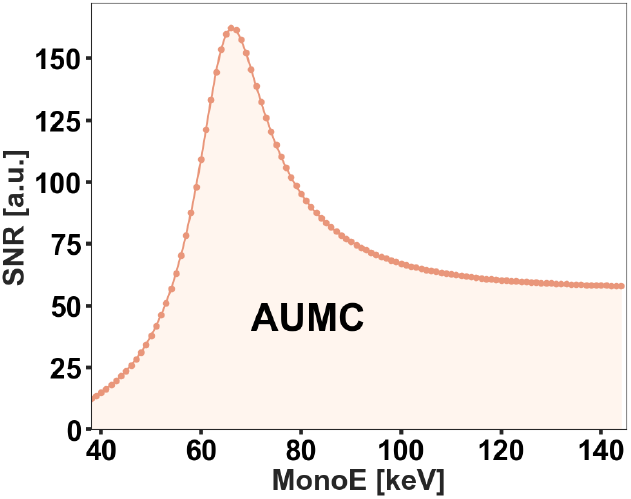
Plot depicting monoenergetic inverse noise values and the AUMC integral area for a kVp-switching system (140/80 kVp, 100 mA, 0.33 duty cycle).

This metric provides a comprehensive assessment of spectral system performance by capturing the cumulative inverse noise across the entire VMI energy spectrum, rather than evaluating image quality at a single energy level. Because AUMC is used here for relative comparisons among different system configurations, its values are reported in arbitrary units.

#### 2.3.4 Dose calculation

In our simulation, dose is estimated by air kerma, which measures the kinetic energy transferred from the photon beam to charged particles per unit mass. Dose evaluation was based on the total photon energy per kVp switching cycle. The simulation uses a voltage-dependent tube emission model specifying the number of photons emitted per solid angle, unit time, and tube current to generate tube spectra *φ*_s_(*V, E*). The tube current itself is also voltage dependent and can be estimated assuming a constant filament temperature of the cathode. The voltage transition times depend on the emission current and can be estimated accordingly. With the specified tube voltages, duty cycle, and transition times, the voltage and current waveforms *V*(*t*), *I*(*V*(*t*)) can be estimated. Since we are interested in the dose, the effective beam filtration within the tube, additional system filtration, and especially the bowtie and K-edge filtration, are summarized with

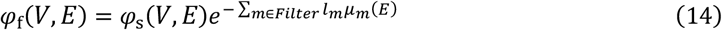

The total photon energy *D* per switching cycle becomes

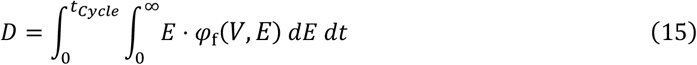

### 2.4 Double bowtie design

The present design considers only the orthogonal projection, with the beam incident perpendicularly to the bed.

#### 2.4.1 Traditional bowtie design - only Teflon

A reference system with the traditional bowtie under reference settings (110/70 kVp, 100 mA, 10/90 duty cycle) serves as a baseline. The bowtie thickness was assumed to be 0 mm at the center ray, and three conventional Teflon bowtie filters, one for each patient size, were designed to maintain a uniform non-spectral detector SNR across all fan angles. The estimated patient radiation exposure profile was determined by attenuating the X-ray beam spectrum according to the Teflon path length at each fan angle. The resulting simulated dose and performance evaluations from the Teflon bowtie filter at given fan beam angles served as reference values for evaluating the double bowtie.

#### 2.4.2 Center optimization - only K-edge material

The double bowtie design started from the center because no Teflon was assumed there, allowing the impact of K-edge materials to be evaluated independently. Non-radioactive and accessible K-edge materials, gadolinium (Gd), holmium (Ho), erbium (Er), silver (Ag), and tin (Sn), were tested at varying filter thicknesses less than 1 mm. Due to the additional attenuation introduced by K-edge materials, tube parameters (tube current and duty cycle) were adjusted to ensure the patient dose remained consistent with the reference system. Tube current was maximally limited by X-ray generator power (80 kW) while the duty cycle was limited by the speed of switching mechanisms and a minimum integration period width. For each K-edge material thickness under the adjusted parameters, updated AUMC as well as *SNR*_40 *keV*_ and *SNR*_70 *keV*_ were calculated and compared to the reference value without K-edge filtration.

Since both K-edge thickness and tube power were adjusted simultaneously, it was essential to isolate the contribution of K-edge materials to performance improvement. Titanium (Ti), which lacks a K-edge in this energy regime, served as a control material. Once the optimal K-edge material and system settings were determined, Ti was tested at varying thicknesses under the same scan parameters until it produced the dose equivalent to the reference. This ensured any observed performance enhancement was attributed solely to the K-edge effect rather than increased tube efficiency.

#### 2.4.3 Full fan-angle optimization - combination of Teflon and K-edge material

The thickness and system settings that provided the highest relative AUMC were selected for the full fan angle filter design. *Flat double-bowtie:* For the simplest configuration, a flat K-edge material filter with uniform optimized thickness across all fan angles was used. Due to the fan beam geometry, the path length through the flat K-edge material increased toward the edges, leading to higher attenuation. The Teflon path length was optimized to ensure the dose profile matched the reference system. After updating the filter profile, the SNR was calculated for monoenergetic images at 40 keV and 70 keV, along with the overall system performance metric, AUMC.

*Fully-optimized double-bowtie:* Based on this, the thickness of both the Teflon bowtie and the K-edge material filter was adjusted simultaneously to achieve an optimal performance across all fan angles. The objective was to maximize the AUMC value while maintaining dose consistency with the reference system. The Scipy Nelder-Mead method in Python was used to match the dose in every fan angle.

## 3. RESULTS

For the reference system, the generated reference bowties create various dose and AUMC profiles for each patient size **(Figure 5A)**. At the center ray, increasing K-edge material thickness led to higher tube current and duty cycle to compensate for increased attenuation. Tube current reached the maximum allowed at 0.28 mm for Gd and Ho, and at 0.29 mm for Er. Beyond these thicknesses, tube current remained at its peak value of 727 mA, and further dose compensation was achieved by increasing the duty cycle (**Figure 5B**). The AUMC initially increased with filter thickness but eventually decreased as tube power became limited. The highest AUMC values were achieved at 0.47 mm for Gd and 0.43 mm for both Ho and Er. These peak values occurred at duty cycles of 0.26 for Gd and Ho, and 0.25 for Er. Importantly, the total dose remained constant and matched the reference across all tested configurations.

**Figure 5.**
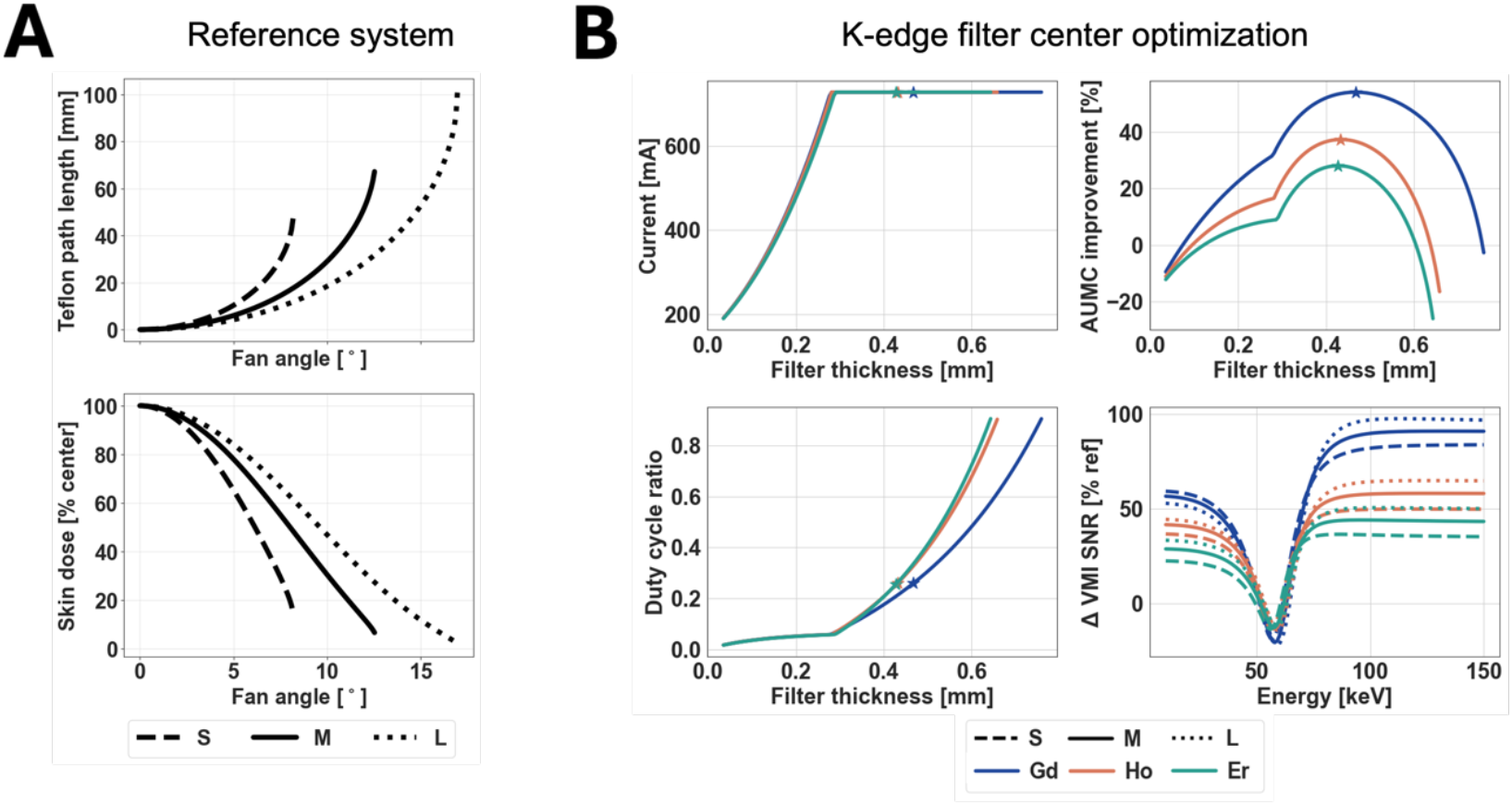
**(A)** Traditional Teflon bowtie profile and dose. *Top:* Teflon bowtie path length at every fan angle for three phantom sizes. *Bottom:* Phantom dose profile compared to the center for each phantom size. **(B)** Center filter optimization procedure. *Top Left:* Adjusted tube current to maintain the required photon fluence through the filter as its thickness increases. *Bottom left:* Duty cycle ratio for the high-kVp tube voltage, adjusted in response to increasing filter thickness. *Top right:* AUMC values across varying filter thicknesses for three K-edge materials, compared to the reference at the center. The filter thickness at which each material achieved its highest AUMC is marked with a star. *Bottom right:* SNR of mono energies relative to the reference system with optimal K-edge filter thickness. Line styles represent phantom sizes, and color represents different filter materials.

For the central ray, the SNR at each VMI energy level using the optimal K-edge filter was calculated, and the relative improvements are presented in **Figure 5B**. Reference SNR values were defined separately for each phantom size. Initially, the relative SNR improvement was positive but declined and dropped to below zero, reaching a minimum around 55 keV. In the energy range between 50 and 63 keV, differences in improvement between phantom sizes and materials were minimal. Beyond this range, the positive SNR improvement increased steadily with energy, eventually reaching a plateau: approximately 90% for Gd, 60% for Ho, and 42% for Er. Among the phantoms, the largest showed the greatest overall improvement, with differences of up to 7% compared to the smallest phantom.

**Figure 6** shows the improvement of both K-edge and non-K-edge compared to the performance of the reference system at the center. The best AUMC was achieved using a Gd thickness of 0.467 mm for the medium and large phantoms, and 0.463 mm for the small phantom. Tube currents reached the highest possible value (727 mA) under tube power limitation for tests of all K-edge materials. For AUMC, Gd was the best K-edge material among all materials for all phantom sizes, having 53.8% average improvement in three phantom sizes. Ho and Er also increased the AUMC by 37.1% and 27.9% on average. Ag and Sn performed similarly, having relatively stable AUMC improvement of around 21% for all phantom sizes. Ti, which lacks a K-edge, reduced AUMC by roughly 6.8% when used at the center. For all materials, larger phantoms had more improvement than smaller ones compared to the corresponding reference system.

**Figure 6.**
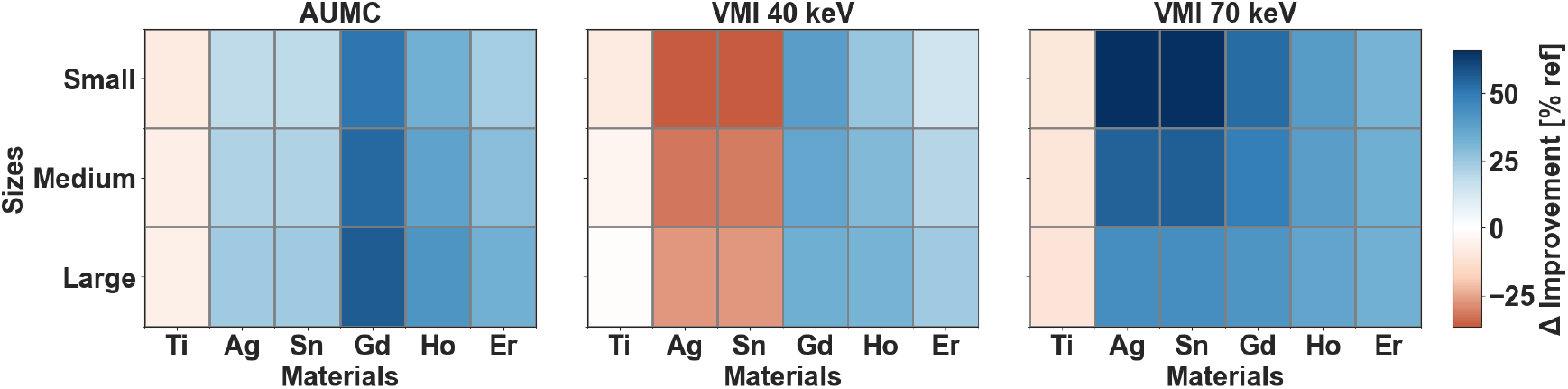
System performance under the highest AUMC conditions for different K-edge materials. The *left* panel shows maximum AUMC values relative to the reference across phantom sizes; the *middle* and *right* panels display corresponding SNR improvements at 40 keV and 70 keV, respectively.

Ho and Er show comparable performance at both 40 keV and 70 keV, with less then 10% difference for each energy level. Ag and Sn had approximately 55% improvements in 70 keV SNR, which were the highest among all K-edge materials, but the 40 keV SNRs were 32% worse than the reference. The suboptimal performance of Ag and Sn can be attributed to their K-edge positions (25 keV and 29 keV). In contrast, Gd provided consistently strong improvements at both 40 keV and 70 keV, making it the most balanced performer across the spectrum.

The flat K-edge filter double bowtie design incorporated a K-edge filter with uniform thickness, which was determined based on center-ray optimization. The three best-performing materials, Gd, Ho, and Er, were selected for full fan-angle evaluation. Due to the fan beam geometry, the effective path length through the K-edge filter increased with fan angle. To maintain dose consistency, the Teflon bowtie was slightly modified from the reference geometry, resulting in path lengths marginally higher (less than 5%) than those in the reference system. Most of the additional attenuation introduced by the K-edge layer was compensated by adjustments to the tube current and duty cycle.

All three K-edge materials showed strong AUMC performance across the full fan range. Gd achieved the highest overall improvement, with an average AUMC increase of 47.51%, followed by Ho at 34.89% and Er at 26.69% **(Table 1)**. The maximum improvement occurred at the center, where the patient thickness was largest. As the fan angle increased, AUMC gradually decreased, with Gd showing the steepest drop (17.78%) from center to edge. Ti, used as a non-K-edge control, was tested under the same tube settings as Gd with a calibrated thickness matching Gd’s attenuation profile. While the Ti filter showed a slight AUMC gain of ~5% at the center, performance declined rapidly beyond 5% and ultimately dropped by 11.34% at the phantom edge. With the comparison between Gd and Ti, the K-edge material showed a contribution to AUMC improvement isolated from tube current increases.

**Table 1.** Performance evaluation of double bowtie with flat K-edge filter for medium phantom over all fan angles.

| | ΔSNR improvement [% ref] | | | | ΔAUMC improvement [% ref] | |
| --- | --- | --- | --- | --- | --- | --- |
|  | VMI 40 keV |  | VMI 70 keV |  |  |  |
|  | Average | Min / Max | Average | Min / Max | Average | Min / Max |
| Teflon + Ti | -2.3 | -4.1 / 0.7 | -10.0 | -10.3 / -9.5 | -6.0 | -6.9 / -4.6 |
| Teflon + Gd | 27.3 | 12.7 / 17 | 38.4 | 22.8 / 49.1 | 47.5 | 36.9 / 54.6 |
| Teflon + Ho | 27.5 | 22.4 / 30.6 | 33.8 | 25.0 / 39.8 | 34.9 | 30.2 / 38.0 |
| Teflon + Er | 19.7 | 17.7 / 20.9 | 29.4 | 22.8 / 33.8 | 26.7 | 23.7 / 28.7 |

The flat double–bowtie design served as a baseline for further optimization, in which the path lengths of the Teflon and K-edge filters were refined. For Ho, Er, and Ti, their path lengths were lower than the previous optimized value at the center, and Teflon’s path length increased (**Figure 7**). The relative AUMC improvement had a similar trend with a flat double bowtie. Specially, the improvement has an increase at the edge for Gd, and at the center for Er. The difference in AUMC improvement was within 5% overall path length for all three K-edge materials. As a comparison, the flat double bowtie has a performance of 13% within optimized double bowtie configurations on average for Gd, Ho, and Er, proving that it achieved similar performance improvement with simple geometry. No matter the geometry, the Ti double bowtie always performed worse than the K-edge material double bowtie designs.

**Figure 7.**
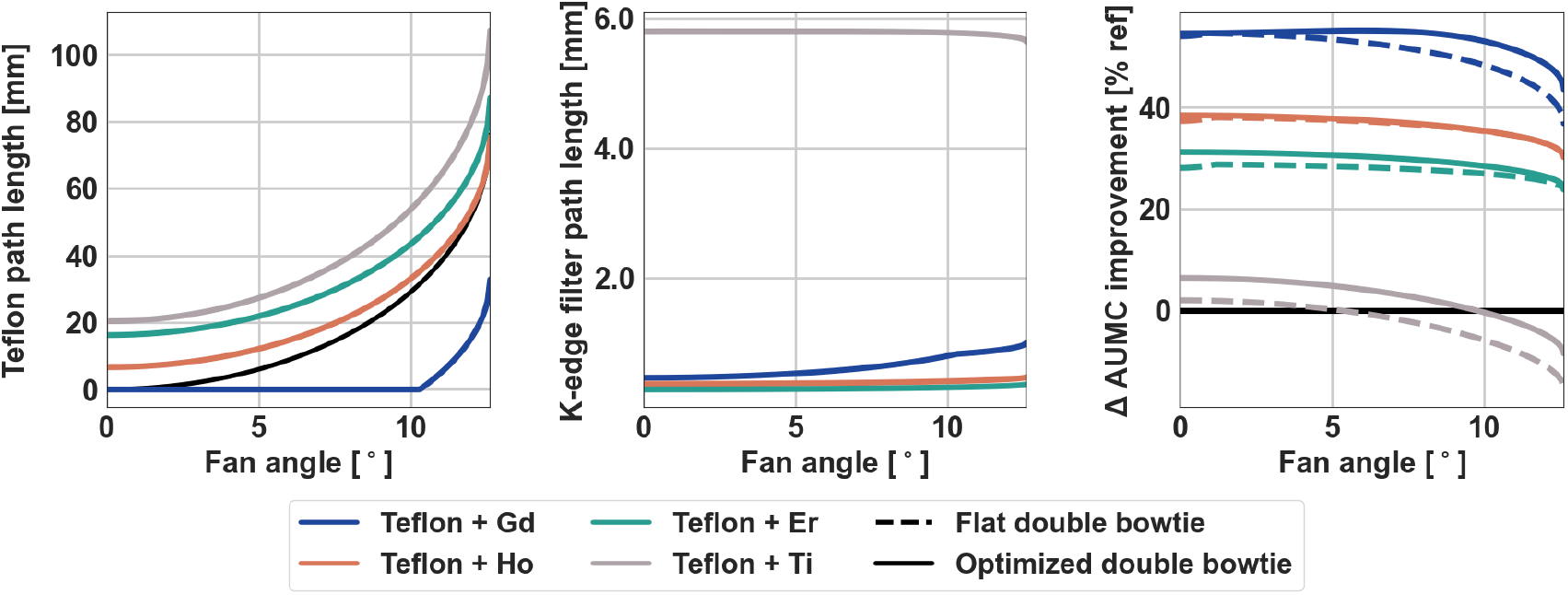
Double bowtie design with fully optimized K-edge filter for medium phantom size. *Left*: Path length of Teflon bowtie. *Middle*: Path length of filters as the ray passes through. *Right*: The AUMC improvement is compared to the reference. The black solid line represents reference bowtie (no K-edge filter). The solid lines with color represent the fully optimized double bowtie, and dashed lines represent the double bowtie with a flat K-edge filter.

Since the flat double bowtie profile was designed, it can be used as a baseline for further optimization, where the path length of Teflon and K-edge filters was optimized. For Ho, Er, and Ti, their path lengths were lower than the previous optimized value at the center, and Teflon’s path length increased (**Figure 7**). The relative AUMC improvement had a similar trend with a flat double bowtie. Specially, the improvement has an increase at the edge for Gd, and at the center for Er. The difference in AUMC improvement was within 5% overall path length for all three K-edge materials. As a comparison, the flat double bowtie has a performance of 13% within optimized double bowtie configurations on average for Gd, Ho, and Er, proving that it achieved similar performance improvement with simple geometry. Regardless of geometry, the Ti double–bowtie consistently underperformed compared with the K-edge-based designs.

As Teflon + Gd was the best combination in the fully optimized double–bowtie configuration, it was further optimized for three phantom sizes (small, medium, and large). Table 2 presents the percentage improvements in VMI 40 keV, VMI 70 keV, and AUMC across these phantoms relative to their respective references. For VMI 40 keV, the average SNR improvement remained consistent across sizes, with values of 49.3%, 47.2%, and 45.6% for small, medium, and large phantoms. At VMI 70 keV, the smaller phantom also had higher improvement. While the small phantom achieved the highest peak value (up to 54%), the medium phantom showed the greatest variability.

**Table 2.** Performance evaluation of Teflon + Gd fully optimized double bowtie for different phantom sizes.

| Phantom | ΔVMI 40 keV improvement [%ref] | | ΔVMI 70 keV improvement [%ref] | | ΔAUMC improvement [%ref] | |
| --- | --- | --- | --- | --- | --- | --- |
|  | Average (%) | Min / Max (%) | Average (%) | Min / Max (%) | Average (%) | Min / Max (%) |
| Small | 49.3 | 39.4 / 55.4 | 41.8 | 35.1 / 53.6 | 47.9 | 45.0 / 51.7 |
| Medium | 47.2 | 36.8 / 57.2 | 37.4 | 22.1 / 49.2 | 51.2 | 43.6 / 55.2 |
| Large | 45.6 | 33.9 / 58.7 | 34.1 | 8.4 / 41.8 | 56.0 | 41.3 / 58.0 |

For AUMC, the improvement trend rose with increasing phantom size, yielding average gains of 47.9%, 51.2%, and 56.0% for small, medium, and large phantoms, respectively. The standard deviation for the medium phantom was higher than that of the small and large phantoms. The maximum AUMC improvement occurred for the large phantom (58%).

## 4. DISCUSSION

A novel prefiltration architecture, termed the double bowtie, was developed to enhance spectral imaging performance in pediatric CT without increasing radiation dose. The design integrates a K-edge material layer with a conventional Teflon bowtie, yielding substantial improvements in spectral signal-to-noise ratio across the VMI range. Among the materials evaluated, gadolinium exhibited the most consistent and robust enhancement.

The performance of the double bowtie depends strongly on the interaction between K-edge material properties and tube operating parameters. When the K-edge material and tube settings were optimized concurrently, both tube current and duty cycle increased with greater filter thickness. Higher tube current resulted in shorter voltage transition durations, while an elevated duty cycle extended the duration of high tube voltage, thereby increasing total energy output. Collectively, these effects enhanced spectral separation by minimizing intermediate kVp intervals and enabling more aggressive duty cycles with shorter integration periods. Titanium, employed as a non–K-edge reference under identical tube conditions, confirmed that the observed improvement arose from K-edge attenuation rather than tube parameter variations. Two filter geometries were examined for full fan-beam optimization: a flat K-edge filter and a thickness-optimized configuration. The flat design achieved the highest AUMC improvement at the central ray, whereas the thickness-optimized filter yielded more uniform SNR enhancement across all fan angles. Despite these geometric differences, their overall performance was comparable, indicating that the simpler flat K-edge filter represents a practical and manufacturable alternative. This simplicity is advantageous for clinical translation, where fabrication efficiency is a key consideration. Furthermore, the high attenuation of K-edge materials allows for substantial reductions in total filter thickness, conserving space within the collimation assembly.

The effectiveness of K-edge filtration is inherently spectrum-dependent, as these materials preferably attenuate the overlapping regions between low- and high-energy spectra. Consequently, the optimal K-edge filter varies with CT system design and tube voltage pairings. Prior studies have demonstrated that holmium performs best in dual-layer detector systems, whereas gadolinium yields superior results in kVp-switching configurations^34^. The 110/70 kVp pair used in this work aligns well with typical pediatric protocols, providing an effective balance between dose efficiency and spectral separation.

Yao et al. ^45^ demonstrated the feasibility of a K-edge prefiltration design for spectral CT using simulation, reporting up to a twofold increase in spectral SNR through optimized K-edge layering under fixed-dose conditions. Wang et al. ^46^also showed the potential of spectral optimization using fast kV switching and filtration to reduce noise in material decomposition. The present work extends the concept by (1) systematically comparing multiple K-edge materials, (2) introducing the AUMC metric to quantify spectral performance across the full energy range, and (3) evaluating manufacturable geometries—both flat and thickness-optimized—within a full fan-beam configuration. Additional approaches to enable spectral CT through K-edge filtration have included the use of commercially available split-beam systems^9,47^ and experimental prototypes incorporating dynamically controlled or spatially varying multi-material filters^48–50^. However, such designs rely on complex hardware architectures that are less compatible with clinical implementation in pediatric imaging, where simplicity, stability, and manufacturability are critical.

Several limitations should be acknowledged. Although optimized bowtie profiles were generated for three pediatric phantom sizes, clinical implementation must accommodate a broader range of patient anatomies and potential size mismatches. Off-center patient positioning, which frequently occurs in practice, may also affect dose uniformity and image quality^51^. Additionally, the increased tube current and duty cycle required for optimal operation demand higher tube power and energy consumption, potentially raising concerns about system power limits and tube longevity. The present design is therefore most applicable to pediatric imaging, where lower tube power settings are standard, and may require modification for adult imaging protocols. Finally, the radiation dose was estimated using air kerma as a surrogate. Although air kerma and absorbed photon energy were shown to exhibit similar spatial distributions, more clinically relevant indices, such as CTDI_vol_, would provide a more accurate estimate of patient exposure. Future work will include advanced simulations and benchtop experiments to evaluate the impact of the double-bowtie design on image quality and its potential for integration into clinical spectral CT systems.

## 5. CONCLUSION

In this study, we present a bowtie filter design integrated with spectral CT system modulation to enhance spectral performance while maintaining low radiation dose levels suitable for pediatric imaging. The design combines a conventional Teflon bowtie with a K-edge material layer optimized across discrete fan angles to improve spectral separation. By incorporating a strategically selected K-edge component, the filter selectively shapes the X-ray spectrum, thereby increasing energy separation and enhancing spectral SNR. Although the current implementation is demonstrated within a kVp-switching CT system, the double-bowtie concept is broadly applicable to other spectral CT technologies and does not require dedicated pediatric hardware. As a pediatric-specific adaptation, this design enables children to benefit fully from the diagnostic advantages of spectral imaging.

## Data Availability

All data produced in the present work are contained in the manuscript

## ACKNOWLEDGEMENTS

We acknowledge support through the National Institutes of Health (R01EB030494) and Philips Healthcare.

## REFERENCES

1. Taguchi K, Blevis I, Iniewski K. Spectral, Photon Counting Computed Tomography: Technology and Applications. 1st ed. (Taguchi K, Blevis I, Iniewski K, eds.). CRC Press; 2020. doi:10.1201/9780429486111

2. Parakh A, Lennartz S, An C, et al. Dual-Energy CT Images: Pearls and Pitfalls. RadioGraphics. 2021;41(1):98–119. doi:10.1148/rg.2021200102

3. Baturin P, Alivov Y, Molloi S. Spectral CT imaging of vulnerable plaque with two independent biomarkers. Phys Med Biol. 2012;57(13):4117–4138. doi:10.1088/0031-9155/57/13/4117

4. Tao S, Rajendran K, Zhou W, Fletcher JG, McCollough CH, Leng S. Improving iodine contrast to noise ratio using virtual monoenergetic imaging and prior-knowledge-aware iterative denoising (mono-PKAID). Phys Med Biol. 2019;64(10):105014. doi:10.1088/1361-6560/ab17fa

5. Michalak G, Grimes J, Fletcher J, et al. Technical Note: Improved CT number stability across patient size using dual-energy CT virtual monoenergetic imaging: CT number stability using dual-energy virtual monoenergetic imaging. Med Phys. 2016;43(1):513–517. doi:10.1118/1.4939128

6. Cester D, Eberhard M, Alkadhi H, Euler A. Virtual monoenergetic images from dual-energy CT: systematic assessment of task-based image quality performance. Quant Imaging Med Surg. 2022;12(1):726–741. doi:10.21037/qims-21-477

7. Gallo-Bernal S, Peña-Trujillo V, Gee MS. Dual-energy computed tomography: pediatric considerations. Pediatr Radiol. 2024;54(13):2112–2126. doi:10.1007/s00247-024-06074-5

8. Li B, Yadava G, Hsieh J, Chandra N, Kulpins MS. Head and body CTDI w of dual-energy x-ray CT with fast-kVp switching. In: 2010:76221Y. doi:10.1117/12.844314

9. Euler A, Parakh A, Falkowski AL, et al. Initial Results of a Single-Source Dual-Energy Computed Tomography Technique Using a Split-Filter: Assessment of Image Quality, Radiation Dose, and Accuracy of Dual-Energy Applications in an In Vitro and In Vivo Study. Invest Radiol. 2016;51(8):491–498. doi:10.1097/RLI.0000000000000257

10. Siegel MJ, Bhalla S, Cullinane M. Dual-Energy CT Material Decomposition in Pediatric Thoracic Oncology. Radiol Imaging Cancer. 2021;3(1):e200097. doi:10.1148/rycan.2021200097

11. So A, Nicolaou S. Spectral Computed Tomography: Fundamental Principles and Recent Developments. Korean J Radiol. 2021;22(1):86. doi:10.3348/kjr.2020.0144

12. Liu LP, Shapira N, Halliburton SS, et al. Spectral performance evaluation of a second-generation spectral detector CT. J Appl Clin Med Phys. 2024;25(4):e14300. doi:10.1002/acm2.14300

13. Greffier J, Villani N, Defez D, Dabli D, Si-Mohamed S. Spectral CT imaging: Technical principles of dual-energy CT and multi-energy photon-counting CT. Diagn Interv Imaging. 2023;104(4):167–177. doi:10.1016/j.diii.2022.11.003

14. Stayman JW, Tivnan M, Wang W, Shapira N, Gang GJ, Noël PB. Spectral CT using a fine grid structure and varying x-ray incidence angle. Med Phys. 2021;48(10):6412–6420. doi:10.1002/mp.14853

15. Sellerer T, Noël PB, Patino M, et al. Dual-energy CT: a phantom comparison of different platforms for abdominal imaging. Eur Radiol. 2018;28(7):2745–2755. doi:10.1007/s00330-017-5238-5

16. Krauss B, Grant KL, Schmidt BT, Flohr TG. The Importance of Spectral Separation: An Assessment of Dual-Energy Spectral Separation for Quantitative Ability and Dose Efficiency. Invest Radiol. 2015;50(2):114–118. doi:10.1097/RLI.0000000000000109

17. Karmazyn B, Liang Y, Klahr P, Jennings SG. Effect of Tube Voltage on CT Noise Levels in Different Phantom Sizes. Am J Roentgenol. 2013;200(5):1001–1005. doi:10.2214/AJR.12.9828

18. Wellenberg RHH, Ahmed R, Müller FC, et al. Quantitative evaluation of the effects of dual-energy CT acquisition, reconstruction and postprocessing parameters on virtual Non-Calcium (VNCa) images. Eur J Radiol. 2025;182:111818. doi:10.1016/j.ejrad.2024.111818

19. Borges AP, Antunes C, Curvo-Semedo L. Pros and Cons of Dual-Energy CT Systems: “One Does Not Fit All.” Tomography. 2023;9(1):195–216. doi:10.3390/tomography9010017

20. Tivnan M, Tilley S, Stayman JW. Physical modeling and performance of spatial-spectral filters for CT material decomposition. In: Bosmans H, Chen GH, Gilat Schmidt T, eds. Medical Imaging 2019: Physics of Medical Imaging. SPIE; 2019:45. doi:10.1117/12.2513481

21. Rakvongthai Y, Worstell W, El Fakhri G, Bian J, Lorsakul A, Ouyang J. Spectral CT Using Multiple Balanced K-Edge Filters. IEEE Trans Med Imaging. 2015;34(3):740–747. doi:10.1109/TMI.2014.2358561

22. Boone JM. Method for evaluating bow tie filter angle-dependent attenuation in CT: Theory and simulation results. Med Phys. 2010;37(1):40–48. doi:10.1118/1.3264616

23. Zhang X, Xie J, Su T, et al. Study on the impact of bowtie filter on photon-counting CT imaging. Phys Med Biol. 2024;69(21):215033. doi:10.1088/1361-6560/ad8858

24. Yang K, Ruan C, Li X, Liu B. Data of CT bow tie filter profiles from three modern CT scanners. Data Brief. 2019;25:104261. doi:10.1016/j.dib.2019.104261

25. Zhang G, Marshall N, Jacobs R, Liu Q, Bosmans H. Bowtie filtration for dedicated cone beam CT of the head and neck: a simulation study. Br J Radiol. 2013;86(1028):20130002. doi:10.1259/bjr.20130002

26. Kontson K, Jennings RJ. Bowtie filters for dedicated breast CT: Analysis of bowtie filter material selection. Med Phys. 2015;42(9):5270–5277. doi:10.1118/1.4928476

27. Gang GJ, Mao A, Wang W, et al. Dynamic fluence field modulation in computed tomography using multiple aperture devices. Phys Med Biol. 2019;64(10):105024. doi:10.1088/1361-6560/ab155e

28. Szczykutowicz TP, Mistretta CA. Design of a digital beam attenuation system for computed tomography: Part I. System design and simulation framework: Design of a digital beam attenuation system. Med Phys. 2013;40(2):021905. doi:10.1118/1.4773879

29. Shunhavanich P, Hsieh SS, Pelc NJ. Fluid-filled dynamic bowtie filter: a feasibility study. In: Hoeschen C, Kontos D, Flohr TG, eds. SPIE Proceedings. Vol 9412. SPIE; 2015:94121L. doi:10.1117/12.2081673

30. Liu F, Wang G, Cong W, Hsieh SS, Pelc NJ. Dynamic bowtie for fan-beam CT. J X-Ray Sci Technol. 2013;21(4):579–590. doi:10.3233/XST-130386

31. Wang H, Yin Z, Jin Y, et al. CT dose minimization using personalized protocol optimization and aggressive bowtie. In: Kontos D, Flohr TG, Lo JY, eds. 2016:978338. doi:10.1117/12.2216570

32. McCollough CH, Boedeker K, Cody D, et al. Principles and applications of multienergy CT: Report of AAPM Task Group 291. Med Phys. 2020;47(7). doi:10.1002/mp.14157

33. Sandvold OF, Ge Y, Proksa R, Noël PB. Double bowtie design for high sensitivity pediatric spectral CT. Conf Proc Int Conf Image Form X-Ray Comput Tomogr. 2024;2024:268–271.

34. Ge Y, Sandvold OF, Perkins AE, Proksa R, Noël PB. High-fidelity prefiltration using a double bowtie design for quantitative low-dose pediatric spectral CT imaging. In: Sabol JM, Abbaszadeh S, Li K, eds. Medical Imaging 2025: Physics of Medical Imaging. SPIE; 2025:4. doi:10.1117/12.3046287

35. Poludniowski G, Landry G, DeBlois F, Evans PM, Verhaegen F. SpekCalc: a program to calculate photon spectra from tungsten anode x-ray tubes. Phys Med Biol. 2009;54(19):N433–N438. doi:10.1088/0031-9155/54/19/n01

36. Seltzer S. XCOM-Photon Cross Sections Database, NIST Standard Reference Database 8. Published online 1987. doi:10.18434/T48G6X

37. Kleinman PL, Strauss KJ, Zurakowski D, Buckley KS, Taylor GA. Patient Size Measured on CT Images as a Function of Age at a Tertiary Care Children’s Hospital. Am J Roentgenol. 2010;194(6):1611–1619. doi:10.2214/AJR.09.3771

38. RE Alvarez, A Macovski. Energy-selective reconstructions in X-ray computerised tomography. Phys Med Biol. 1976;21(5):733–744. doi:10.1088/0031-9155/21/5/002

39. Roessl E, Herrmann C. Cramér–Rao lower bound of basis image noise in multiple-energy x-ray imaging. Phys Med Biol. 2009;54(5):1307–1318. doi:10.1088/0031-9155/54/5/014

40. Kay S, Cuichun Xu. CRLB via the characteristic function with application to the K-distribution. IEEE Trans Aerosp Electron Syst. 2008;44(3):1161–1168. doi:10.1109/TAES.2008.4655371

41. Zhong H, Huang Q, Zheng X, et al. Generation of virtual monoenergetic images at 40 keV of the upper abdomen and image quality evaluation based on generative adversarial networks. BMC Med Imaging. 2024;24(1):151. doi:10.1186/s12880-024-01331-3

42. Ishikawa T, Suzuki S, Katada Y, et al. Evaluation of three-dimensional iterative image reconstruction in virtual monochromatic imaging at 40 kilo-electron volts: phantom and clinical studies to assess the image noise and image quality in comparison with other reconstruction techniques. Br J Radiol. 2020;93(1110):20190675. doi:10.1259/bjr.20190675

43. Tatsugami F, Higaki T, Nakamura Y, Honda Y, Awai K. Dual-energy CT: minimal essentials for radiologists. Jpn J Radiol. 2022;40(6):547–559. doi:10.1007/s11604-021-01233-2

44. Leng S, Yu L, Fletcher JG, McCollough CH. Maximizing Iodine Contrast-to-Noise Ratios in Abdominal CT Imaging through Use of Energy Domain Noise Reduction and Virtual Monoenergetic Dual-Energy CT. Radiology. 2015;276(2):562–570. doi:10.1148/radiol.2015140857

45. Yao Y, Wang AS, Pelc NJ. Efficacy of fixed filtration for rapid kVp-switching dual energy x-ray systems. Med Phys. 2014;41(3):031914. doi:10.1118/1.4866381

46. Wang S, Yang Y, Pal D, et al. Spectral optimization using fast kV switching and filtration for photon counting CT with realistic detector responses: a simulation study. J Med Imaging. 2024;11(S1). doi:10.1117/1.JMI.11.S1.S12805

47. Rutt B, Fenster A. Split-Filter Computed Tomography: A Simple Technique for Dual Energy Scanning. J Comput Assist Tomogr. 1980;4(4):501–509. doi:10.1097/00004728-198008000-00019

48. Stayman JW, Tivnan M, Wang W, Shapira N, Gang GJ, Noël PB. Spectral CT using a fine grid structure and varying x-ray incidence angle. Med Phys. 2021;48(10):6412–6420. doi:10.1002/mp.14853

49. Tivnan M, Wang W, Stayman JW. A prototype spatial–spectral CT system for material decomposition with energy-integrating detectors. Med Phys. 2021;48(10):6401–6411. doi:10.1002/mp.14930

50. Xi Y, Cong W, Harrison D, Wang G. Grating Oriented Line-Wise Filtration (GOLF) for Dual-Energy X-ray CT. Sens Imaging. 2017;18(1):27. doi:10.1007/s11220-017-0174-7

51. Gomez-Cardona D, Cruz-Bastida JP, Li K, Budde A, Hsieh J, Chen GH. Impact of bowtie filter and object position on the two-dimensional noise power spectrum of a clinical MDCT system: Symmetry of 2D NPS of MDCT systems. Med Phys. 2016;43(8Part1):4495–4506. doi:10.1118/1.4954848

